# Circulating Cell-Free RNA in Blood as a Host Response Biomarker for the Detection of Tuberculosis

**DOI:** 10.1101/2023.01.11.23284433

**Authors:** Adrienne Chang, Conor J. Loy, Joan S. Lenz, Amy Steadman, Alfred Andama, Nguyen Viet Nhung, Charles Yu, William Worodria, Claudia M. Denkinger, Payam Nahid, Adithya Cattamanchi, Iwijn De Vlaminck

## Abstract

Tuberculosis (TB) remains a leading cause of death from an infectious disease worldwide. This is partly due to a lack of tools to effectively screen and triage individuals with potential TB. Whole blood RNA signatures have been extensively studied as potential biomarkers for TB, but they have failed to meet the World Health Organization’s (WHOs) target product profiles (TPPs) for a non-sputum triage or diagnostic test. In this study, we investigated the utility of plasma cell-free RNA (cfRNA) as a host response biomarker for TB. We used RNA profiling by sequencing to analyze plasma samples from 182 individuals with a cough lasting at least two weeks, who were seen at outpatient clinics in Uganda, Vietnam, and the Philippines. Of these individuals, 100 were diagnosed with microbiologically-confirmed TB. Our analysis of the plasma cfRNA transcriptome revealed 541 differentially abundant genes, the top 150 of which were used to train 15 machine learning models. The highest performing model led to a 9-gene signature that had a diagnostic accuracy of 89.1% (95% CI: 83.6-93.4%) and an area under the curve of 0.934 (95% CI: 0.8674-1) for microbiologically-confirmed TB. This 9-gene signature exceeds the optimal WHO TPPs for a TB triage test (sensitivity: 96.2% [95% CI: 80.9-100%], specificity: 89.7% [95% CI: 72.4-100%]) and was robust to differences in sample collection, geographic location, and HIV status. Overall, our results demonstrate the utility of plasma cfRNA for the detection of TB and suggest the potential for a point-of-care, gene expression-based assay to aid in early detection of TB.

**One Sentence Summary:** This study is the first to investigate the utility of circulating RNA in plasma as a new class of host response signature for tuberculosis and provides evidence that plasma RNA signatures are highly specific for TB and robust against differences in patient cohorts and sample processing.

## INTRODUCTION

The outcome of infection after exposure to *Mycobacterium tuberculosis*, the causative agent of tuberculosis (TB), can vary greatly and is determined by a complex interplay between host immunity and bacterial persistence^1^. Current diagnostic tests for TB are not sensitive to the full spectrum of disease states, are unable to determine if an infection has been cleared, cannot distinguish between latent, incipient, and subclinical disease, or predict progression to active TB^2,3^. This lack of sensitivity and specificity in diagnostic tests presents a major challenge in the management and control of TB.

Transcriptomic signatures, or changes in host-cell gene expression, can provide valuable insight into the host response to TB. Yet, while several whole blood RNA (wbRNA) signatures have been identified, none have met the target product profiles (TPPs) recommended by the Word Health Organization (WHO) for a non-sputum-based triage or diagnostic test^4–6^.

In this study, we investigated plasma cell-free RNA (cfRNA) as a potential new class of host biomarkers for TB. cfRNA is an analyte that can provide information beyond what can be achieved with conventional wbRNA signatures for two key reasons. First, cfRNA is released by cells in both the blood compartment and from vascularized solid tissues, and thus contains information about systemic immune dynamics and immune-tissue interactions. Second, wbRNA is predominantly derived from live cells, whereas cfRNA is primarily released by dying cells. Thus, cfRNA may provide insights into pathways of cell death and mechanisms of cellular injury that would not be available with wbRNA profiling.

To evaluate the performance of plasma cfRNA as a host signature of TB, we conducted unbiased profiling of plasma cfRNA from 182 individuals with cough ≥ 2 weeks enrolled in two independent clinical studies in Uganda, Vietnam, and the Philippines. Of these individuals, 100 were diagnosed with microbiologically confirmed TB. Our analysis identified panels of differentially abundant genes, which we used to train 15 machine learning models. Our top performing model revealed a 9-gene signature that has a diagnostic accuracy of 89.1%, exceeds the optimal WHO TPPs for a triage test (sensitivity: 0.962 and specificity: 0.897), and is robust against differences in sample collection, HIV status, and geographic area. These results provide the foundation for the development and clinical validation of a point-of-care, gene expression-based assay for the early detection of TB.

## RESULTS

### Plasma cell-free RNA signatures of tuberculosis

We analyzed plasma samples from 182 individuals with a cough lasting at least two weeks who were enrolled in two independent clinical studies (END TB and R2D2) at outpatient clinics in Uganda, Vietnam, and the Philippines (**Table 1** and **Figure 1A**). Individuals included in the “TB positive” group were required to have 1) a positive Xpert MTB/RIF Ultra on sputum, urine, or contaminated Mycobacterial Growth Indicator Tube (MGIT) specimen; 2) a positive sputum MGIT or solid culture; or, 3) two trace Xpert Ultra results on sputum or contaminated MGIT. All other individuals (“TB negative” group) had at least one negative Xpert Ultra result and two negative cultures in MGIT or solid media. Of the 182 plasma samples analyzed in our study, 100 (56%) were from individuals diagnosed with microbiologically-confirmed TB and 159 (87%) were from HIV-negative individuals. We used next-generation sequencing to profile the cfRNA in these plasma samples. This yielded a mean of 33,016,810 reads per sample (range: 11,176,564 to 57,039,590 reads; **Figure S1**).

**Table 1:** Patient Information

|  | END TB |  | R2D2 |  |
| --- | --- | --- | --- | --- |
|  | TB Positive | TB Negative | TB Positive | TB Negative |
| n | 53 | 36 | 47 | 46 |
| Age, median (IQR) | 29 (25-35) | 30 (24.75-38.25) | 47.5 (32-57.75) | 27 (38-52) |
| Females, n (%) | 14 (26.4) | 20 (55.6) | 16 (34.0) | 35 (50.0) |
| HIV positive, n (%) | 6 (11.3) | 8 (22.2) | 1 (0.02) | 8 (17.4) |
| Prior TB, n (%) | 4 (7.5) | 9 (25) | 11 (23.4) | 4 (8.7) |

**Figure 1.**
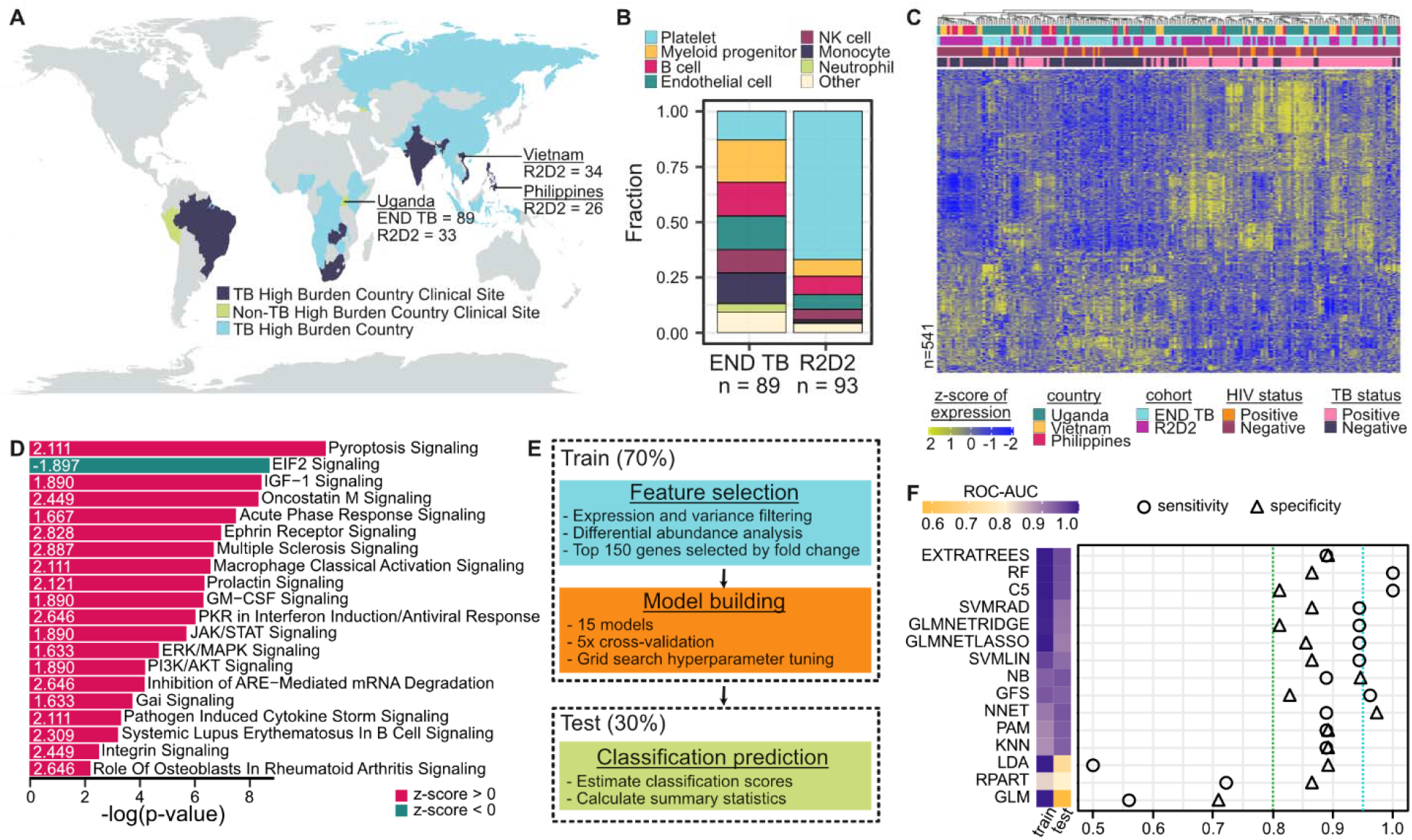
Plasma cell-free RNA profiling. **A)** Geographic distribution of samples included in this study, which originated from two clinical studies. **B)** Differences in cfRNA cell-types-of-origin between the two cohorts. **C)** Scaled counts per million values of significantly differentially abundant genes after batch correction (DESeq2 and limma, Benjamini-Hochberg adjusted p-value < 0.01, |Log2FoldChange| > 0.5). Samples and genes are clustered based on correlation. **D)** Top 20 differential pathways between microbiologically confirmed TB diagnoses ranked by significance. Z-score indicated in corresponding bar. **E)** Flowchart of the method used to train and test machine learning classification algorithms. **F)** (Left) Area under the receiver operating characteristic curve (ROC-AUC) metrics for training and test sets. (Right) The test sensitivity and specificities for each model. The dotted lines indicate the optimal sensitivity (blue) and specificity (green) for a triage test.

To determine the cell-types-of-origin of the cfRNA in our samples, we used a reference-based deconvolution algorithm (BayesPrism). This algorithm uses a reference dataset, in this case the Tabula Sapiens single-cell RNA-seq atlas, and bayesian inference to identify the cell types from which the cfRNA in our samples originated^7–10^. We found that the plasma cfRNA in individuals with respiratory symptoms was primarily derived from platelets, myeloid progenitor cells, B cells, endothelial cells, natural killer cells, monocytes, and neutrophils (**Figure 1B**). We observed batch effects in the platelet contribution, with a mean fraction of 0.13 and 0.67 in the END TB and R2D2 cohorts, respectively (Mann-Whitney U adjusted p-value=4.7 × 10^−31^). These differences were due to the different plasma collection procedures used by the END TB and R2D2 studies (Methods).

To gain insight into the host response to TB and to identify potential diagnostic biomarkers, we performed differential abundance analysis using the R package DESeq2^11^. Using a log2 fold change cutoff of 0.5 and a Benjamini-Hochberg adjusted p-value of <0.01, we identified 541 differentially abundant genes between TB positive and negative groups (**Figure 1C** and **Supplementary File**). TB was associated with elevated levels of macrophage markers (*MARCO, SOCS3, FCGR1A, MPO, C1QB*), neutrophil markers (*ELANE, FCGR3A, S100A8, S100A9, ERG*), interferon genes (*IFI27L1, IFIT2, IFIT3, IFITM3, IRF1*), and antimicrobial genes (*AZU1, CTSG, DEFA4, STAT1, GBP1, GBP2, GBP4, GBP5, GBP7*) compared to the TB negative group. We also observed elevated levels of lung-specific markers (*SFTPC, SFTPB, AGER, SLC34A2, SFTPA1, SFTPA2, RTKN2*) in individuals with TB, providing insight into ongoing pathogenic processes^12,13^. To confirm the relevance of these findings, we performed pathway analysis using Ingenuity Pathway Analysis (**Figure 1D**). The top canonical pathways enriched in TB (ranked by -log(p-value) > 1.5 and |z-score|>1.3) included pyroptosis signaling, macrophage classical activation signaling, and pathogen-induced cytokine storm signaling pathways.

### Evaluating machine learning algorithms for TB diagnosis

The results from the differential expression analysis, including enrichment of clinically relevant pathways and satisfactory separation of sample groups using correlation-based clustering, suggested that differentially abundant genes in plasma cfRNA could be used to develop triage tests and diagnostic assays for TB. To test this idea, we randomly divided the samples into a training set (70%) and a test set (30%), controlling for cohort and disease status (**Figure 1E**). We then performed feature selection on the training set by first removing genes that were poorly detected, had low counts, or had near zero variance (Methods). Next, we performed differential abundance analysis using DESeq2 with batch-corrected gene counts (limma removeBatchEffect) and selected the top 150 relevant features (Benjamini-Hochberg adjusted p-value < 0.01, ranked by absolute fold change). The raw counts of these top 150 genes were then normalized (CPM) and used to train 14 machine learning classification models as described in Sotomayor-Gonzalez et al.^14^ and a greedy forward search algorithm as described in Sweeney et al^15^. Classification score thresholds were determined by Youden’s J statistic and used to predict the disease status of samples in the test set (Methods).

The majority of the models performed well, with 10/15 models having a train and test area under the receiver operating characteristic curve (ROC-AUC) > 0.9 (**Figure 1F** and **Table S1**). Decision tree algorithms produced the top three models: greedy forward search, C5.0, and random forest. These models had test ROC-AUCs of >0.93 and test specificities and sensitivities exceeding the optimal thresholds for a triage test (95% sensitivity, 80% specificity). However, some models, such as the generalized linear model (GLM) and linear discriminant analysis (LDA), suffered from overfitting, as they had perfect training performance but poor test performance. The poor performance of GLM may be due to the lack of regularization, since the performance of the generalized linear models with Ridge and LASSO feature selection (GLMNETRIDGE and GLMNETLASSO) were on par with the other models, suggesting that there may a number of highly correlated features. This high collinearity may also explain the poor performance of LDA.

### 9-gene biomarker panel surpasses guidelines for a non-sputum-based triage test

After evaluating the performance of the different machine learning models, we based the candidate biomarker panel off the greedy forward search results because it was the most consistent and highest performing model across training and test sets. The greedy forward search algorithm begins by choosing the gene with the most discriminatory power in terms of a TB score, which is calculated as the mean expression of the downregulated genes subtracted from the mean expression of the upregulated genes. In each subsequent iteration, the algorithm adds the gene that produces the greatest increase in ROC-AUC. This yielded a 9-gene signature consisting of guanylate binding protein 5 (GBP5), dysferlin (DYSF), SWI/SNF related, matrix associated, actin dependent regulator of chromatin, subfamily D, member 3 (SMARCD3), vesicle associated membrane protein 5 (VAMP5), guanylate binding protein 2 (GBP2), Fc gamma receptor 3B (FCGR3B), glucose-6-phosphate isomerase (GPI), myeloperoxidase (MPO), and CAMP responsive element binding protein 5 (CREB5) (**Figure 2A** and **Table S2**).

**Figure 2.**
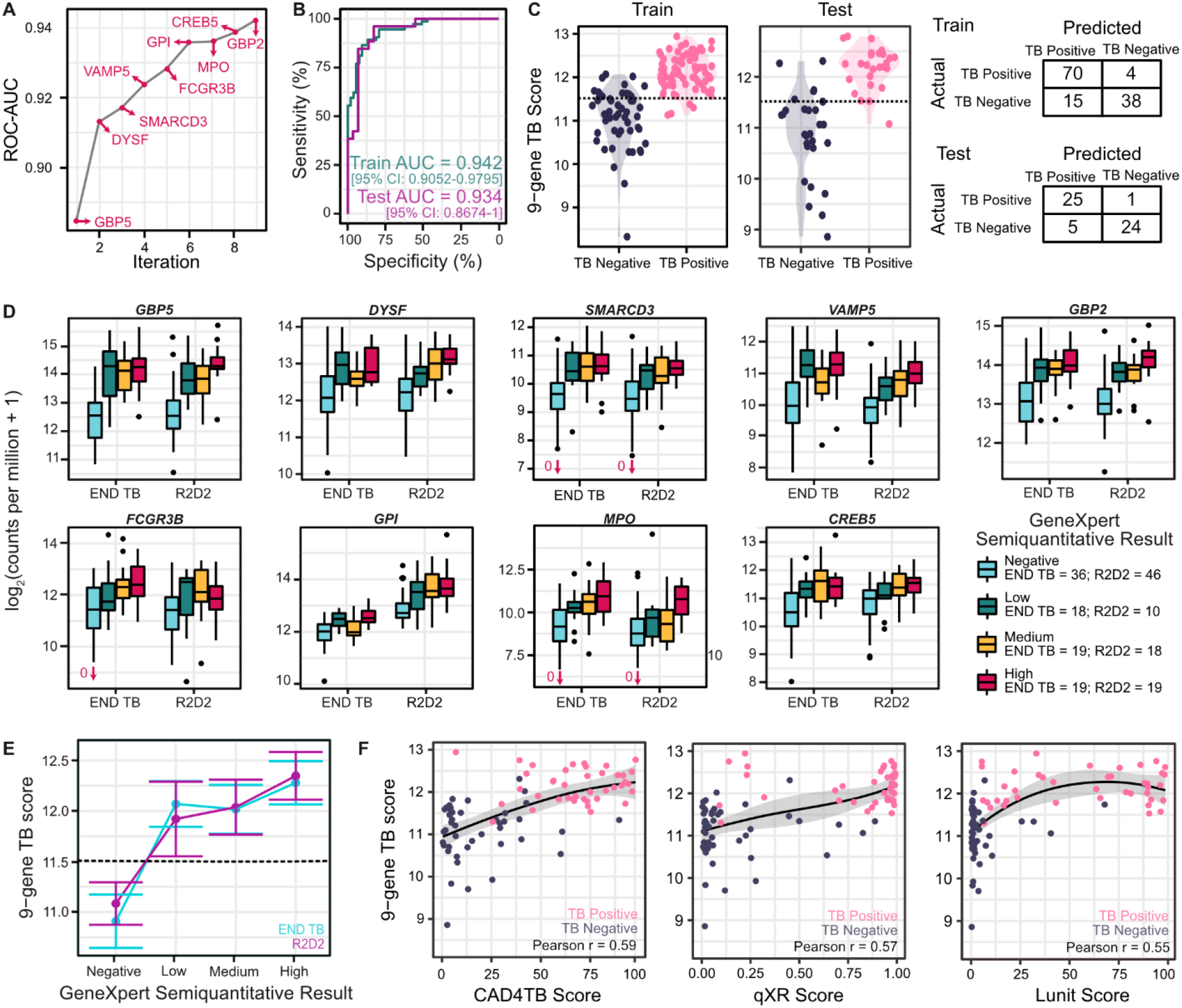
Performance of a 9-gene signature identified in plasma cfRNA. **A)** Test area under the receiver operating characteristic curve (ROC-AUC) as a function of the gene added in each iteration of a greedy forward search model. **B)** Train and test performance of the greedy forward search algorithm in distinguishing microbiologically confirmed TB. **C)** Violin plot of classifier scores and confusion matrices of training and test sets using the greedy forward search algorithm. **D)** Abundance of the genes included in the 9-gene TB score. Outliers are indicated with arrows and values. **E)** Correlation of a 9-gene TB score with the Xpert Ultra Semi-quantitative Results (dotted line: classification score threshold=11.5; bars indicate 95% confidence interval). **F)** Correlation of a 9-gene TB score with three chest X-ray scores.

We evaluated the utility of the 9-gene signature as a diagnostic or triage tool using ROC analysis on the training and test sets. The WHO TPPs provide the minimal and optimal characteristics of a target product, where the minimal thresholds indicate the lowest acceptable standards and the optimal criteria outline the ideal target profile. For a non-sputum-based diagnostic test, the overall minimal and optimal sensitivities are 80% and 65% with a specificity of 98%; for a non-sputum-based triage test, the optimal requirements are 95% sensitivity and 80% specificity, while the minimal criteria are 90% sensitivity and 70% specificity^6^. We found that the discriminatory power of the signature was high in both sets (training set ROC-AUC=0.942 vs test set ROC-AUC=0.934; **Figure 2B**). In the test set, the TB score discriminated between TB positive and negative groups with 89.1% accuracy, 96.2% sensitivity, and 89.7% specificity (**Figure 2C** and **Table S3**). The mean TB score for TB positive and TB negative individuals was 12.14 and 11.0, respectively (adjusted p-value=2.9 × 10^−24^, Wilcoxon test). Although the biomarker panel falls short of the specificity requirements for a diagnostic test, it exceeds the optimal criteria for a triage test (optimal: 95% sensitivity, 80% specificity; minimal: 90% sensitivity, 70% specificity). We found that the discriminatory power of the 9-gene signature was independent of geographic area, HIV status, and cohort. When we evaluated each gene individually, we found that, except for *GPI*, the normalized counts of each of the genes were robust against differences in sample collection (**Figure 2D** and **Table S4**). Furthermore, the TB score was highly correlated with the Xpert Ultra Semiquantitative result, which bins samples based on the cycle threshold (CT) of the first positive probe that detects *M. tuberculosis* as follows: “low” = 22<CT≤38, “medium” = 16<CT≤22, and “high” = CT≤16 (polyserial correlation=0.8213; **Figure 2E** and **Tables S5-S6**). These results indicate that the 9-gene signature has the potential to serve as a valuable triage tool for TB.

Finally, we compared the 9-gene signature to chest X-ray (CXR) scores, as radiographs show evidence of the host response to TB. Recent developments in computer-aided detection (CAD) software have improved the performance of CXRs, but still fall short of the WHO TPPs for a triage test^16^. Three commercially available CAD applications were used to analyze CXRs from individuals enrolled in the R2D2 study: CAD4TB (Delft Imaging, Netherlands), qXR (Quire.ai, India), and Lunit Insight chest X-ray TB algorithm (Lunit Inc., South Korea). The TB abnormality score is reported on a scale of 0-100 (for CAD4TB and Lunit) or 0-1 (for qXR), where a higher value indicates a more abnormal radiograph. Despite performance differences between each of the CAD algorithms, we find that the 9-gene TB score is moderately correlated with the reported TB abnormality scores (Pearson r=0.55-0.59; **Figure 2F**), indicating that the gene expression signature may provide complementary information to chest X-ray results in the evaluation of TB.

### Comparison of plasma cfRNA biomarkers to published whole blood RNA, protein assays

While hundreds of potential wbRNA signatures have been identified for the diagnosis of TB disease in clinically-relevant populations, only a handful meet the minimal WHO criteria for a triage test and none meet the minimal WHO criteria for a diagnostic test^4,17,18^. Unsurprisingly, the performance of these wbRNA signatures varies widely when applied to multi-country cohorts, as diagnostic performance heterogeneity between populations is a well-known barrier to the development of triage and diagnostic assays^19^.

We found that 7 of the 9 genes in our signature overlap with those reported in previous wbRNA studies (**Figure 3A**)^4^. Among the wbRNA signatures that meet the minimal WHO criteria for a triage test, 7 have been evaluated for their performance in distinguishing TB from other diseases in a multi-cohort population (Berry86^20^, daCosta2^21^, daCosta3^21^, Kaforou44^22^, Walter47^23^, Zak16^5^, and Sweeney3^15^; **Figure 3B**). The performance of these 7 signatures varies widely, with sensitivities and specificities ranging from 79.3-100% and 80-95%, respectively. Collectively, the 7 wbRNA signature gene sets and our 9-gene signature gene set include 178 genes, of which 166 are unique to a single signature set.

**Figure 3.**
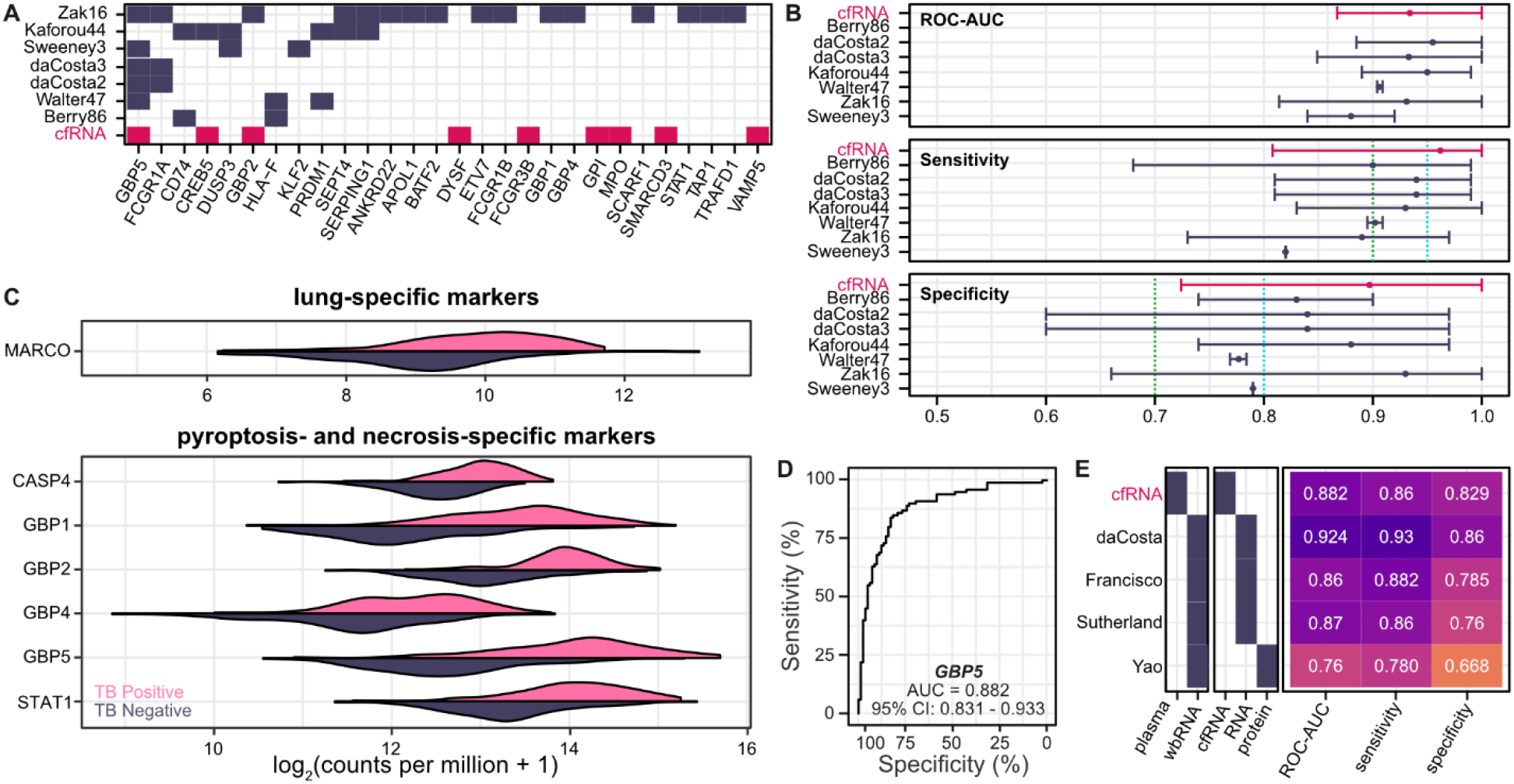
Comparison of whole blood and plasma signatures of TB. **A)** Overlap between top-performing whole blood signatures (blue) and the 9-gene signature (pink). Top 28 overlapping genes are shown. **B)** Performance comparison of whole blood signatures (blue) with the 9-gene signature (pink). Optimal triage thresholds are marked in blue, minimal triage thresholds are marked in green. **C)** Abundance of lung-specific markers non-programmed cell-death markers. **D)** Performance of *GBP5* mRNA abundance in distinguishing active TB. **E)** Performance comparison of whole blood protein GBP5, whole blood RNA *GBP5*, and plasma cfRNA *GBP5* in distinguishing active TB.

To further evaluate the differences between plasma cfRNA and wbRNA in relation to TB, we examined tissue-specific markers that would provide evidence of a host response. We identified elevated lung-, pyroptosis-, and necrosis-specific markers in TB positive individuals in the top 150 differentially expressed genes using the Human Protein Atlas and the Tabula Sapiens dataset^10,24^. These include macrophage receptor with collagenous structure (*MARCO*), caspase 4 (*CASP4*), members of the guanylate binding protein family (*GBP1, GBP2, GBP4, GBP5*), and signal transducer and activator of transcription 1 (*STAT1*) (**Figure 3C** and **Table S7**).

*GBP5* is the most frequently occurring transcript, occurring in the 9-gene signature and in 5/7 whole blood signatures. *GBP5* has been shown to be critical for NLRP3 inflammasome activation, which mediates caspase-1 activation and secretion of proinflammatory cytokines in response to pathogenic bacteria and cellular damage^25,26^. The discriminatory power of *GBP5* alone has been investigated in three of the wbRNA studies: da Costa et al.^21^ report a sensitivity of 93%, specificity of 86%, and ROC-AUC of 0.924; Francisco et al.^27^ report a sensitivity of 88.2%, specificity of 78.5%, and ROC-AUC of 0.86; and Sutherland et al.^28^ (Xpert-MTB-HR prototype using the Sweeney3^15^ signature) reports a sensitivity of 86%, specificity of 76%, and an ROC-AUC of 0.87. In comparison, we find that the abundance of *GBP5* in plasma results in a sensitivity of 86%, specificity of 82.9%, and an ROC-AUC of 0.882 (**Figures 3D-E**). Whole blood GBP5 protein levels have also been evaluated as a potential biomarker for TB. Yao et al. demonstrate that whole blood GBP5 protein levels are significantly higher in people with microbiologically-confirmed TB^29^. However, the performance of the plasma protein biomarker at the optimal threshold (sensitivity=0.7802, specificity=0.6691, ROC-AUC=0.76) is poorer than the plasma RNA biomarker. This suggests that plasma cfRNA is a more effective biomarker for TB than whole blood GBP5 protein levels.

## DISCUSSION

In this study, we investigated the use of plasma cfRNA as a host-specific biomarker for the detection of TB. Using RNA sequencing and machine learning, we developed a 9-gene signature that can accurately distinguish individuals with microbiologically diagnosed TB. Our signature has a diagnostic accuracy of 89.1%, sensitivity of 96.2%, and specificity of 89.7%, which compares favorably to the performance of previous whole blood signatures. This performance meets the World Health Organization’s (WHOs) target product profile (TPP) for a TB triage test, making it a promising candidate for further development. This is particularly important because non-sputum-based biomarker tests are a priority for TB management, as many high-risk populations are unable to produce high-quality sputum samples for diagnosis.

Our study is the first to investigate the potential of plasma cfRNA as a host-specific blood-borne biomarker for TB. Given that plasma cfRNA is a novel bioanalyte to monitor TB, these results are important in at least three ways. First, our results suggest that plasma cfRNA signatures may be more robust than whole blood RNA (wbRNA) signatures, which have shown poor performance in independent validation studies^4,18^. This may be due to the sensitivity of whole blood signatures to differences in patient cohorts and sample processing. We found that the plasma cfRNA signature is robust against these factors, making it a potentially more reliable biomarker for TB. Second, plasma cfRNA is stable and can be assayed using a small amount of plasma, making it a practical option for point-of-care diagnostics. Finally, future studies could explore the use of combined wbRNA and cfRNA bioanalytes for the development of more sensitive and specific diagnostic assays for TB.

Our study has several limitations that should be considered when interpreting the results. The most important limitation is that our study does not include an independent validation cohort, so further validation is needed to ascertain the clinical utility of our cfRNA signatures. Additionally, our study does not include analysis of matched whole blood samples to facilitate a full analysis of blood compartment differences in gene expression. Despite these limitations, we demonstrate that the majority of the genes included in our 9-gene signature are robust against batch effects and overlap with those described in whole blood transcriptional signatures. These observations suggest that plasma cfRNA has the capability to serve as a proxy for tissue-specific changes in gene expression and may more adequately capture the host response to TB than wbRNA. Further research is needed to confirm these findings and develop a point-of-care, gene expression-based assay based on the plasma cfRNA signature. Such an assay could be a valuable tool in the early detection of TB and help improve the management and control of this disease.

## MATERIALS AND METHODS

### Ethics statement

All patients provided written informed consent and all experiments were performed in accordance with relevant guidelines and regulations. The protocols for this study were approved locally at each site by Institutional Review Boards at Cornell University (protocols IRB0145569, 1902008555); UCSF (protocol 20-32670); Heidelberg University (S-539/2020); the Makerere University School of Medicine (protocol 2017-020); Vietnam National Lung Hospital (protocol 566/2020/NCKH), and De La Salle Medical and Health Sciences Institute (protocol 2020-33-02-A).

### Sample collection

Plasma samples were collected as part of two independent clinical studies: Evaluation of Novel Diagnostics for Tuberculosis (END TB) and the Rapid Research in Diagnostics Development for TB Network (R2D2). The studies enrolled adults with cough ≥2 weeks identified at outpatient clinics in Uganda (END TB) and in Uganda, Vietnam, and the Philippines (R2D2).

Peripheral blood samples from individuals enrolled in the END TB study in Uganda were collected in Streck Cell-Free DNA blood collection tubes (Streck, 230257). Plasma was separated by centrifugation at 1,600 *x g* for 10 minutes at ambient temperature. The remaining cellular debris was removed by an additional centrifugation step at 16,000 *x g* for 10 minutes at ambient temperature. Plasma was stored in 1 mL aliquots at -80°C.

Plasma samples from individuals enrolled in the R2D2 study sites in Uganda, Vietnam, and the Philippines were collected in K2EDTA blood collection tubes (BD Diagnostics, 366643). Plasma was separated by centrifugation at 1,600 *x g* for 10 minutes at ambient temperature in a horizontal rotor (swing out head). Plasma was similarly stored in 1 mL aliquots at -80°C.

### cfRNA isolation and library preparation

Plasma samples were received on dry ice and stored at -80°C until processed. Prior to cfRNA extraction, plasma samples were thawed at room temperature and centrifuged at 1300 *x g* for 10 minutes at 4°C. cfRNA was extracted from plasma (300-1,000 µl) using the Norgen Plasma/Serum Circulating and Exosomal RNA Purification Mini Kit (Norgen, 51000). Extracted RNA was DNase treated with 14 µl of 10 µl DNase Turbo Buffer (Invitrogen, AM2238), 3 µl DNase Turbo (Invitrogen, AM2238), and 1 µl Baseline Zero DNase (Lucigen-Epicenter, DB0715K) for 30 minutes at 37°C, then concentrated into 12 µl using the Zymo RNA Clean and Concentrated Kit (Zymo, R1015).

Sequencing libraries were prepared from 8 µl of concentrated RNA using the Takara SMARTer® Stranded Total RNA-Seq Kit v3 - Pico Input Mammalian (Takara, 634485) and barcoded using the SMARTer® RNA Unique Dual Index Kit (Takara, 634451). Library concentration was quantified using the Qubit™ 3.0 Fluorometer (Invitrogen, Q33216) with the dsDNA HS Assay Kit (Invitrogen, Q32854). Libraries were quality-controlled using the Agilent Fragment Analyzer 5200 (Agilent, M5310AA) with the HS NGS Fragment Kit (Agilent, DNF-474-0500) and pooled to equal concentrations. Each pool was sequenced using both the Illumina NextSeq 500/550 platform (paired-end, 150 bp) and the Illumina NextSeq 2000 platform (paired-end, 100 bp).

### Bioinformatic processing and sample quality filtering

Sequencing data was processed using a custom bioinformatics pipeline. Since pools sequenced on the Illumina NextSeq 2000 were optimized to produce paired-end, 61 bp reads, matched sequencing data from the Illumina NextSeq 500 were trimmed using Seqtk (v1.2). Samples were then quality filtered and trimmed using BBDuk (v38.90), aligned to the Gencode GRCh38 human reference genome (v38, primary assembly) using STAR^30^ (v2.7.0f, default parameters). Prior to feature quantification using featureCounts^31^ (v2.0.0), samples were deduplicated using Picard MarkDuplicates (v2.19.2). Mitochondrial, ribosomal, X, and Y chromosome genes were bioinformatically removed prior to analysis.

Samples were filtered on the basis of DNA contamination, rRNA contamination, total counts, and RNA degradation. DNA contamination was estimated by calculating the ratio of reads mapping to introns and exons. rRNA contamination was measured using SAMtools (v1.14). Total counts were calculated using featureCounts^31^ (v2.0.0). Degradation was estimated by calculating the 5’-3’ bias using Qualimap^32^ (v2.2.1).

Samples were removed from analysis if either the intron to exon ratio was greater than 3, if a sample had less than 75,000 total counts, or if the rRNA contamination, total counts, or 5’-3’ bias was greater than three standard deviations from the mean.

### Cell type deconvolution

Cell type deconvolution was performed using BayesPrism^9^ (v1.1) with the Tabula Sapiens single-cell RNA-seq atlas^10^ (Release 1) as a reference. Cells from the Tabula Sapiens atlas were grouped as previously described in Loy et al^7^. Cell types with more than 100,000 unique molecular identifiers (UMIs) were included in the reference and subsampled to 300 cells using ScanPy^33^ (v1.8.1).

### Differential abundance analysis

Comparative analysis of differentially expressed genes was performed using a negative binomial model as implemented in DESeq2^11^ (v1.34.0). Heatmaps were constructed using the pheatmap package in R (v1.0.12). Samples and genes were clustered using correlation-based hierarchical clustering. Canonical pathways, diseases and functions were analyzed using QIAGEN Ingenuity Pathway Analysis software (v73620684).

### Machine learning and model training

Machine learning and model training was performed using R (v4.1.3) with the DESeq2^11^ (v1.34.0), Caret (v6.0.90), limma (v3.50.3), and pROC^34^ (v1.18.0) packages. Sample metadata and count matrices were split 70/30 into a training set and a test set. Cohort and disease status were considered while splitting the data to minimize differences between the training and test sets.

Features for model training were selected by filtering and differential abundance analysis. First, genes having low counts (sum counts per million across samples < 20), near zero variance (R caret package nearZeroVar function), or poorly detected genes (less than 0.5 CPM in at least 75% of training samples) were removed. Following this step, we retained 25,947 genes for differential abundance analysis. Differentially abundant genes were identified using DESeq2^11^ (using HIV status and cohort as covariates), after batch correction (R limma package removeBatchEffect) to account for differences in sample collection methods, and ranked by absolute log_2_ fold change (adjusted p-value < 0.01). The top 150 genes were selected for model training.

Machine learning algorithms were trained using 5-fold cross validation and grid search hyperparameter tuning. Accuracy, sensitivity, specificity, and area under the receiver operating characteristic curve (ROC-AUC) were used to measure test performance. The classification models used were generalized linear models with Ridge and LASSO feature selection (GLMNETRIDGE and GLMNETLASSO), support vector machines with linear and radial basis function kernel (SVMLin and SVMRAD), random forest (RF), random forest ExtraTrees (EXTRATREES), neural networks (NNET), linear discriminant analysis (LDA), nearest shrunken centroids (PAM), C5.0 (C5), k-nearest neighbors (KNN), naive bayes (NB), CART (RPART), and logistic regression (GLM).

An additional model was trained and tested using a greedy forward search algorithm (GFS), as described in Sweeney et al^15^. Briefly, the algorithm starts with a single gene that provides the most discriminatory power using a TB score. The TB score was calculated as:

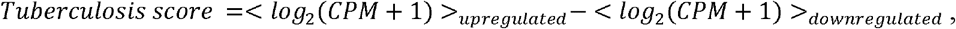

where CPM is counts per million obtained using the cpm function (edgeR v3.36.0) on raw counts. At each subsequent step, an additional gene is added and the TB score is recalculated. The combination of genes at each step that provides the greatest increase in ROC-AUC becomes the basis for the next iteration. The search stops when no gene addition increases the ROC-AUC.

### Quantification and statistical analyses

All statistical analyses were performed using R (v4.1.0). Statistical significance was tested using Wilcoxon signed-rank tests and Mann-Whitney U tests in a two-sided manner, unless otherwise stated. Boxes in the boxplots indicate the 25th and 75th percentiles, the band in the box represents the median, and whiskers extend to 1.5 x interquartile range of the hinge. All sequencing data was aligned to the GRCh38 Gencode v38 Primary Assembly and features were counted using the GRCH38 Gencode v38 Primary Assembly Annotation.

## Data Availability

All code will be made available on Github. Processed sequencing data will be deposited in the National Institutes of Health (NIH) / National Center for Biotechnology Information (NCBI) Sequence Read Archive (SRA) and Gene Expression Omnibus (GEO) repositories under restricted access via Database for Genotypes and Phenotypes (dbGAP).

## List of Supplementary Materials

Fig. S1

Tables S1 to S7

Data file S1

## Acknowledgments

We thank the Cornell Genomics Center and the UCSF Center for Advanced Technology for help with sequencing libraries. We thank the patients and their families for contributing their blood to further our understanding of TB.

## Author contributions

Study design: AC, AS, IDV

Experiments: JSL

Data analysis: AC, CJL, IDV

Data collection: AS, AC, AA, NVN, CY, WW, CMD, PN

Writing – original draft: AC, IDV

Writing – review & editing: AC, CJL, JSL, AS, AA, NVN, CY, WW, CMD, PN, AC, IDV

