## Supplementary Information for "Circulating Cell-Free RNA in Blood as a Host Response Biomarker for the Detection of Tuberculosis"

**Affiliations:** ^1^Meinig School of Biomedical Engineering, Cornell University, Ithaca, NY, USA; ^2^Global Health Labs, Inc. Bellevue, WA; ^3^Walimu, Kampala, Uganda; ^4^National Lung Hospital, Hanoi, Vietnam; ^5^De La Salle Medical and Health Sciences Institute, Dasmarinas, Philippines; ^6^University Hospital Heidelberg & German Center of Infection Research, Heidelberg, Germany; ^7^Center for Tuberculosis, University of California San Francisco, San Francisco, CA, USA; ^8^Division of Pulmonary and Critical Care Medicine, University of California Irvine, Orange, CA, USA

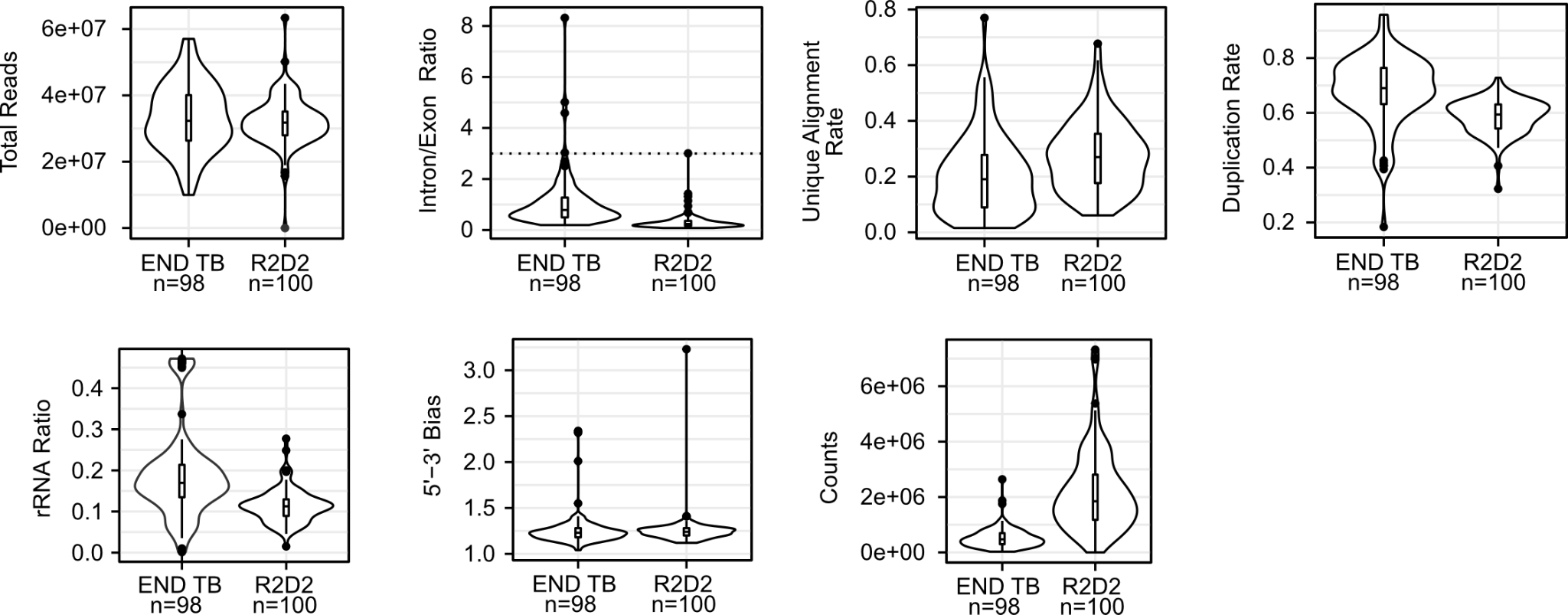

### Figure S1. Quality control metrics used to filter samples. Samples were filtered on the basis of DNA contamination (estimated by the intron/exon ratio), total counts (calculated using featureCounts), rRNA contamination, and RNA degradation (estimated by the 5’-3’ bias).

### Table S1. Performance metrics of the 15 models on the training and test sets.

| **Model** | **Train ROC-AUC**  **[95% CI]** | **Test ROC-AUC [95% CI]** | **Train Accuracy**  **[95% CI]** | **Test Accuracy**  **[95% CI]** | **Train Sensitivity**  **[95% CI]** | **Test Sensitivity [95% CI]** | **Train Specificity**  **[95% CI]** | **Test Specificity [95% CI]** |
| --- | --- | --- | --- | --- | --- | --- | --- | --- |
| C5 | 1.00  [1.00-1.00] | 0.946  [0.893-0.999] | 1.00  [0.95-1.00] | 0.645  [0.454-0.808] | 1.00  [1.00-1.00] | 1.00  [0.833-1.00] | 1.00  [1.00-1.00] | 0.811  [0.803-0973] |
| EXTRATREES | 1.00  [1.00-1.00] | 0.952  [0.901-1.00] | 1.00  [0.95-1.00] | 0.839  [0.663-0.945] | 1.00  [1.00-1.00] | 0.889  [0.722-0.944] | 1.00  [1.00-1.00] | 0.892  [0.676-1.00] |
| GLM | 1.00  [1.00-1.00] | 0.574  [0.426-0.723] | 1.00  [0.95-1.00] | 0.636 [0.4910.764] | 1.00  [1.00-1.00] | 0.56  [0.0-0.89] | 1.00  [1.00-1.00] | 0.703  [0.432-1.00] |
| GLMNETLasso | 0.999 [0.996-1.00] | 0.905 [0.829-0.982] | 0.992 [0.976-1.00] | 0.855 [0.745-0.927] | 0.984 [0.953-1.00] | 0.944 [0.722-1.00] | 1.00 [1.00-1.00] | 0.855 [0.745-0.927] |
| GLMNETRidge | 0.993 [0.98-1.00] | 0.913 [0.838-0.988] | 0.976 [0.953-1.00] | 0.855 [0.745-0.95] | 0.984 [0.922-1.00] | 0.944  [0.722-0.944] | 0.984 [0.937-1.00] | 0.811 [0.649-0.973] |
| NNET | 0.911 [0.856-0.965] | 0.944 [0.879-1.00] | 0.882 [0.827-0.929] | 0.927 [0.818-0.982] | 0.859 [0.766-0.95] | 0.889 [0.667-1.00] | 0.905 [0.810-0.984] | 0.973 [0.784-1.00] |
| RF | 1.00  [1.00-1.00] | 0.95 [0.90-1.00] | 1.00  [0.95-1.00] | 0.891 [0.782-0.964] | 1.00  [1.00-1.00] | 1.00  [0.722-1.00] | 1.00  [1.00-1.00] | 0.865 [0.676-1.00] |
| SVMLin | 0.962 [0.831-0.993] | 0.923 [0.854-0.993] | 0.921 [0.974-0.961] | 0.873 [0.764-0.946] | 0.891 [0.781-0.969] | 0.944 [0.722-1.00] | 0.968 [0.857-1.00] | 0.865 [0.649-1.00] |
| LDA | 1.00  [1.00-1.00] | 0.709  [0.556-0.861] | 1.00  [0.95-1.00] | 0.806  [0.625-0.925] | 1.00  [1.00-1.00] | 0.5  [0.333-0.611] | 1.00  [1.00-1.00] | 0.892  [0.351-0.973] |
| SVMRAD | 0.994 [0.987-1.00] | 0.916 [0.844-0.988] | 0.969 [0.945-0.992] | 0.873 [0.745-0.945] | 0.984 [0.891-1.00] | 0.944 [0.778-1.00] | 0.984 [0.889-1.00] | 0.865 [0.622-0.973] |
| PAM | 0.896  [0.85-0.954] | 0.938  [0.868-1.00] | 0.847  [0.743-0.921] | 0.742  [0.554-0.881] | 0.859  [0.75-0.938] | 0.889  [0.772-0.889] | 0.857  [0.778-0.952] | 0.892  [0.811-1.00] |
| KNN | 0.890 [0.831-0.949] | 0.935 [0.869-1.00] | 0.866 [0.803-0.921] | 0.891 [0.782-0.964] | 0.859 [0.734-0.938] | 0.889 [0.667-1.00] | 0.873 [0.778-0.952] | 0.892 [0.703-1.00] |
| RPART | 0.827  [0.762-0.893] | 0.794  [0.673-0.914] | 0.903  [0.810-0.960] | 0.806  [0.625-0.925] | 0.766  [0.656-0.875] | 0.722  [0.5-0.722] | 0.889  [0.810-0.952] | 0.865  [0.757-0.973] |
| NB | 0.934  [0.891-0.978] | 0.944  [0.884-1.00] | 0.890 [0.843-0.937] | 0.909 [0.8-0.982] | 0.891 [0.781-0.984] | 0.889 [0.722-1.00] | 0.905 [0.778-0.968] | 0.946 [0.703-1.00] |
| GFS | 0.942  [0.905-0.980] | 0.934  [0.867-1.00] | 0.882  [0.835-0.937] | 0.891  [0.836-0.964] | 0.865  [0.797-0.973] | 0.962  [0.808-1.00] | 0.906  [0.774-0.981] | 0.828  [0.758-1.00] |

### Table S2. Genes and model performance of each step in the greedy forward search.

| **Step #** | **ENSEMBL ID** | **Gene Name** | **ROC-AUC [95% CI]** |
| --- | --- | --- | --- |
| 1 | ENSG00000154451.14 | *GBP5* | 0.885 [0.827-0.943] |
| 2 | ENSG00000135636.15 | *DYSF* | 0.913 [0.864-0.962] |
| 3 | ENSG00000082014.17 | *SMARCD3* | 0.917 [0.866-0.968] |
| 4 | ENSG00000168899.5 | *VAMP5* | 0.924 [0.874-0.974] |
| 5 | ENSG00000162747.12 | *FCGR3B* | 0.928 [0.886-0.971] |
| 6 | ENSG00000105220.17 | *GPI* | 0.935 [0.895-0.976] |
| 7 | ENSG00000005381.8 | *MPO* | 0.936 [0.894-0.978] |
| 8 | ENSG00000146592.17 | *CREB5* | 0.939 [0.899-0.978] |
| 9 | ENSG00000162645.13 | *GBP2* | 0.942 [0.905-0.980] |

### Table S3. Greedy forward search performance on the test set at various thresholds.

| **Threshold** | **Accuracy [95% CI]** | **Sensitivity [95% CI]** | **Specificity [95% CI]** | **PPV [95% CI]** | **NPV [95% CI]** |
| --- | --- | --- | --- | --- | --- |
| Youden | 0.891  [0.836-0.934] | 0.962  [0.808-1] | 0.897  [0.724-1] | 0.833 [0.765-1] | 0.96  [0.829-1] |
| Triage (min sensitivity) | 0.862  [0.698-0.953] | 0.9 | 0.828  [0.517-1] | 0.824  [0.626-1] | 0.902  [0.852-0.918] |
| Triage (min specificity) | 0.824  [0.769-0.842] | 0.962  [0.846-1] | 0.7 | 0.742  [0.712-0.749] | 0.953  [0.835-1] |
| Diagnostic (min sensitivity) | 0.798  [0.744-0.835] | 0.65 | 0.931  [0.828-1] | 0.894 [0.772-1] | 0.748  [0.725-0.761] |
| Diagnostic (min specificity) | 0.699  [0.626-0.953] | 0.385  [0.231-0.923] | 0.98 | 0.945 [0.912-0.976] | 0.640  [0.5870.934] |

### Table S4. Comparison of normalized counts (CPM) per gene and cohort, Wilcoxon test.

| **Gene** | **Semiquantitative Xpert Ultra result** | **Group 1** | **Group 2** | **Group 1 Mean [95% CI]** | **Group 2 Mean [95% CI]** | **Adjusted p-value** |
| --- | --- | --- | --- | --- | --- | --- |
| GBP5 | Negative | END TB | R2D2 | 12.56 [12.27-12.86] | 12.63 [12.37-12.90] | 0.89 |
|  | Low | END TB | R2D2 | 13.94 [13.40-14.49] | 13.91 [13.30-14.52] | 0.83 |
|  | Medium | END TB | R2D2 | 14.03 [13.68-14.37] | 13.82 [13.45-14.19] | 0.48 |
|  | High | END TB | R2D2 | 14.22 [13.85-14.59] | 14.38 [14.06-14.70] | 0.47 |
| DYSF | Negative | END TB | R2D2 | 12.15 [11.86-14.45] | 12.15 [11.95-12.36] | 0.81 |
|  | Low | END TB | R2D2 | 12.94 [12.66-13.21] | 12.73 [12.39-13.06] | 0.31 |
|  | Medium | END TB | R2D2 | 12.60 [12.39-12.81] | 12.99 [12.71-12.27] | 0.033 |
|  | High | END TB | R2D2 | 12.92 [12.68-13.15] | 13.15 [12.96-13.34] | 0.085 |
| SMARCD3 | Negative | END TB | R2D2 | 8.82 [7.87-9.78] | 8.99 [8.33-9.64] | 0.71 |
|  | Low | END TB | R2D2 | 10.53 [10.15-10.91] | 10.26 [9.77-10.75] | 0.33 |
|  | Medium | END TB | R2D2 | 10.55 [10.15-10.95] | 10.28 [9.89-10.68] | 0.44 |
|  | High | END TB | R2D2 | 10.62 [10.29-10.95] | 10.59 [10.37-10.80] | 0.71 |
| VAMP5 | Negative | END TB | R2D2 | 10.12 [9.78-10.47] | 9.88 [9.67-10.10] | 0.29 |
|  | Low | END TB | R2D2 | 11.28 [10.97-11.60] | 10.55 [10.12-10.98] | 0.0072 |
|  | Medium | END TB | R2D2 | 10.72 [10.32-11.11] | 10.71 [10.36-11.06] | 0.91 |
|  | High | END TB | R2D2 | 11.24 [10.87-11.61] | 11.03 [10.75-11.32] | 0.23 |
| FCGR3B | Negative | END TB | R2D2 | 10.74 [9.78-11.69] | 11.31 [11.02-11.59] | 0.94 |
|  | Low | END TB | R2D2 | 12.00 [11.55-12.44] | 11.80 [10.79-12.81] | 0.72 |
|  | Medium | END TB | R2D2 | 12.36 [11.93-12.79] | 12.12 [11.61-12.62] | 0.72 |
|  | High | END TB | R2D2 | 12.52 [12.13-12.90] | 11.90 [11.55-12.25] | 0.018 |
| GPI | Negative | END TB | R2D2 | 11.97 [11.79-12.14] | 12.88 [12.72-13.03] | 1 x 10^-12^ |
|  | Low | END TB | R2D2 | 12.48 [12.32-12.64] | 13.37 [12.80-13.95] | 0.0025 |
|  | Medium | END TB | R2D2 | 12.11 [11.88-12.34] | 13.72 [13.37-14.07] | 2.7 x 10^-8^ |
|  | High | END TB | R2D2 | 12.58 [12.43-12.74] | 13.76 [13.44-14.09] | 7.9 x 10^-9^ |
| MPO | Negative | END TB | R2D2 | 8.19 [7.11-9.27] | 8.07 [7.21-8.92] | 0.27 |
|  | Low | END TB | R2D2 | 10.25 [9.82-10.68] | 9.81 [8.47-11.18] | 0.099 |
|  | Medium | END TB | R2D2 | 10.43 [9.75-11.11] | 9.49 [8.81-10.16] | 0.042 |
|  | High | END TB | R2D2 | 10.94 [10.40-11.47] | 10.68 [10.19-11.17] | 0.58 |
| CREB5 | Negative | END TB | R2D2 | 10.55 [10.22-10.87] | 10.79 [10.53-11.02] | 0.17 |
|  | Low | END TB | R2D2 | 11.34 [11.09-11.59] | 11.08 [10.62-11.54] | 0.41 |
|  | Medium | END TB | R2D2 | 11.50 [11.10 -11.90] | 11.41 [11.12-11.70] | 0.85 |
|  | High | END TB | R2D2 | 11.45 [11.17-11.74] | 11.51 [11.28-11.74] | 0.56 |
| GBP2 | Negative | END TB | R2D2 | 13.09 [12.85-13.32] | 13.06 [12.87-13.26] | 0.86 |
|  | Low | END TB | R2D2 | 13.87 [13.57-14.16] | 13.77 [13.42-14.11] | 0.62 |
|  | Medium | END TB | R2D2 | 13.85 [13.62-14.09] | 13.81 [13.59-14.03] | 0.55 |
|  | High | END TB | R2D2 | 14.02 [13.79-14.26] | 14.12 [13.88-14.36] | 0.42 |

### Table S5. Comparison of TB score and semiquantitative Xpert Ultra results for all samples, Wilcoxon test.

| **Group 1** | **Group 2** | **Group 1 Mean [95% CI]** | **Group 2 Mean [95% CI]** | **Adjusted p-value** |
| --- | --- | --- | --- | --- |
| Negative | Low | 11.00 [10.84-11.17] | 12.02 [11.84-12.20] | 3.5 x 10^-10^ |
| Negative | Medium | 11.00 [10.84-11.17] | 12.03 [11.88-12.17] | 2.6 x 10^-12^ |
| Negative | High | 11.00 [10.84-11.17] | 12.31 [12.21-12.42] | 2.6 x 10^-12^ |
| Low | Medium | 12.02 [11.84-12.20] | 12.03 [11.88-12.17] | 0.9832 |
| Low | High | 12.02 [11.84-12.20] | 12.31 [12.21-12.42] | 0.0041 |
| Medium | High | 12.03 [11.88-12.17] | 12.31 [12.21-12.42] | 0.0017 |

### Table S6. Comparison of TB score and cohort, Wilcoxon test.

| **Semiquantitative Xpert Ultra result** | **Group 1** | **Group 2** | **Group 1 Mean [95% CI]** | **Group 2 Mean [95% CI]** | **Adjusted p-value** |
| --- | --- | --- | --- | --- | --- |
| Negative | END TB | R2D2 | 10.91 [10.62-11.20] | 11.08 [10.89-11.28] | 0.44 |
| Low | END TB | R2D2 | 12.07 [11.82-12.32] | 11.92 [11.64-12.20] | 0.31 |
| Medium | END TB | R2D2 | 12.02 [11.81-12.22] | 12.04 [11.81-12.26] | 0.82 |
| High | END TB | R2D2 | 12.28 [12.12-12.44] | 12.35 [12.21-12.49] | 0.62 |

### Table S7. Comparison of lung-, pyroptosis-, and necrosis-specific gene abundances (normalized CPM), cohort, and TB status (Wilcoxon test).

| **Gene** | **Group 1** | **Group 2** | **Group 1 Mean**  **[95% CI]** | **Group 2 Mean [95% CI]** | **Adjusted p-value** |
| --- | --- | --- | --- | --- | --- |
| *MARCO* | END TB TB Positive | END TB TB Negative | 10.06 [9.8-10.32] | 7.75 [6.49-9.01] | 4x10^-5^ |
|  | R2D2 TB Positive | R2D2  TB Negative | 9.73 [9.38-10.08] | 7.92 [7.13-8.71] | 1.9x10^-6^ |
|  | R2D2 TB Negative | END TB TB Negative | 7.92 [7.13-8.71] | 7.75 [6.9-9.01] | 0.28 |
|  | R2D2 TB Positive | END TB TB Positive | 9.73 [9.38-10.08] | 10.06 [9.8-10.32] | 0.15 |
| *CASP4* | END TB TB Positive | END TB TB Negative | 12.9 [12.77-13.03] | 12.4 [12.26-12.54] | 2.3x10^-6^ |
|  | R2D2 TB Positive | R2D2  TB Negative | 13.06 [12.96-13.16] | 12.52 [12.37-12.68] | 5x10^-8^ |
|  | R2D2 TB Negative | END TB TB Negative | 12.52 [12.37-12.68] | 12.4 [12.26-12.54] | 0.11 |
|  | R2D2 TB Positive | END TB TB Positive | 13.06 [12.96-13.16] | 12.9 [12.77-13.03] | 0.19 |
| *GBP1* | END TB TB Positive | END TB TB Negative | 13.27 [13.02-13.53] | 12.17 [11.87-12.46] | 4.5x10^-7^ |
|  | R2D2 TB Positive | R2D2  TB Negative | 13.48 [13.27-13.7] | 12.16 [11.9-12.42] | 1.2x10^-11^ |
|  | R2D2 TB Negative | END TB TB Negative | 12.16 [11.9-12.42] | 12.17 [11.87-12.46] | 0.89 |
|  | R2D2 TB Positive | END TB TB Positive | 13.48 [13.27-13.7] | 13.27 [13.02-13.53] | 0.36 |
| *GBP2* | END TB TB Positive | END TB TB Negative | 13.91 [13.76-14.06] | 13.1 [12.87-13.33] | 8.6x10^-8^ |
|  | R2D2 TB Positive | R2D2  TB Negative | 13.93 [13.78-14.07] | 13.06 [12.87-13.26] | 5.4x10^-10^ |
|  | R2D2 TB Negative | END TB TB Negative | 13.06 [12.87-13.26] | 13.1 [12.87-13.33] | 0.81 |
|  | R2D2 TB Positive | END TB TB Positive | 13.93 [13.78-14.07] | 13.91 [13.73-14.06] | 0.84 |
| *GBP4* | END TB TB Positive | END TB TB Negative | 12.12 [11.91-12.32] | 11.37 [11.05-11.69] | 2x10^-4^ |
|  | R2D2 TB Positive | R2D2  TB Negative | 12.38 [12.2-12.56] | 11.67 [11.43-11.91] | 6.3x10^-6^ |
|  | R2D2 TB Negative | END TB TB Negative | 11.67 [11.43-11.91] | 11.37 [11.05-11.69] | 0.19 |
|  | R2D2 TB Positive | END TB TB Positive | 12.38 [12.2-12.56] | 12.12 [11.91-12.32] | 0.053 |
| *GBP5* | END TB TB Positive | END TB TB Negative | 14.04 [13.79-14.3] | 12.6 [12.32-12.88] | 1.5x10^-9^ |
|  | R2D2 TB Positive | R2D2  TB Negative | 14.07 [13.84-14.29] | 12.63 [12.37-12.9] | 1.8x10^-12^ |
|  | R2D2 TB Negative | END TB TB Negative | 12.63 [12.37-12.9] | 12.6 [12.32-12.88] | 0.99 |
|  | R2D2 TB Positive | END TB TB Positive | 14.07 [13.84-14.29] | 14.04 [13.79-14.3] | 0.85 |
| *STAT1* | END TB TB Positive | END TB TB Negative | 13.89 [13.71-14.07] | 13.12 [12.89-13.36] | 1.7x10^-6^ |
|  | R2D2 TB Positive | R2D2  TB Negative | 14.25 [14.07-14.43] | 13.42 [13.21-13.64] | 1.1x10^-8^ |
|  | R2D2 TB Negative | END TB TB Negative | 13.42 [13.21-13.64] | 13.12 [12.89-13.36] | 0.068 |
|  | R2D2 TB Positive | END TB TB Positive | 14.25 [14.07-14.43] | 13.89 [13.71-14.07] | 0.0075 |

**Description of Additional Supplementary Files**

File Name: Supplementary Data 1 (SuppData1.tsv)

Description: Differential abundances between TB positive and TB negative groups.
